# Meeting the needs of people with pulmonary fibrosis: patient perspectives on improving palliative care provision: a qualitative study

**DOI:** 10.64898/2026.09.28.26364170

**Authors:** Caroline Barry, Carole Gardener, Morag Farquhar, Isha Karwa, Andrew Wilson

## Abstract

**Background:** Palliative care is suboptimal in pulmonary fibrosis. Symptom burden is high despite evidence-based interventions, guidelines, and information resources. The application of existing knowledge and methods to optimally manage palliative and end of life care for people with pulmonary fibrosis in the UK has not been realised.

**Aim:** To establish patient views and understanding of how palliative care services and approaches can better meet the needs of people with pulmonary fibrosis. To identify the current barriers and facilitators to receiving palliative care services.

**Design:** Qualitative exploration of patient views via semi-structured interviews and focus groups. Content analysis was conducted.

**Setting/Participants:** The study was conducted from March–August 2024 within an Interstitial Lung Disease Specialist Centre, based in an NHS acute Trust.

**Results:** Two focus groups and four interviews were conducted, involving nine patients. Four overarching themes were identified as to how palliative care can better meet the needs of this group: (1) establishing mechanisms for identifying and addressing palliative care needs; (2) open channels of communication with healthcare professionals; (3) the importance of a palliative care approach within respiratory, community and specialist services; and (4) support for unpaid/family carers.

**Conclusions:** Patients highlighted the importance of person-centred principles and services that adapted to their needs (such as real-time telephone support), rather than services that imposed their own requirements on the patients. Understanding how existing palliative care provision can meet this standard could help improve the palliative and end of life care for people with pulmonary fibrosis.

**Key Statement:** *What is already known about this topic?:*

- Pulmonary fibrosis is associated with significant patient and carer need, and high symptom burden
- Pulmonary fibrosis is associated with poor outcomes relevant to palliative care, with limited use of advance care plans (ACP), high levels of end-stage healthcare utilisation, frequent hospital deaths and poor symptom control.
- There is consensus that palliative care is beneficial for this condition, but the model of care required is not clear.

*What this paper adds?:*

- Patients with pulmonary fibrosis do not readily distinguish between the care they receive from the specialist respiratory services and that which they receive from palliative care services.
- Accessibility of services is of key importance in addressing palliative care needs.
- Person-centred principles are critical to ensure that services reflect the varying information and care needs of this population.

*Implications for Practice, Theory or Policy:*

- A “one size fits all” approach to palliative care is unlikely to meet the needs of people with pulmonary fibrosis.
- Provision for remote real-time telephone or video contact is important for this group who, due to oxygen and other care needs, may find hospital visits challenging.
- More work is needed to understand how palliative care services and approaches can best meet the needs of unpaid/family carers who support people with pulmonary fibrosis.

## Introduction

Progressive fibrotic lung diseases comprise idiopathic and progressive pulmonary fibrosis (hereafter referred to as pulmonary fibrosis: PF). These are incurable, progressive lung conditions with medial survival of 3 to 7 years (1–3) and are associated with significant patient and carer need and high symptom burden, particularly breathlessness, pain and fatigue (4, 5). Known palliative care needs include information provision, support from family/friends, psychological support, symptom relief and medical management, and end of life care.

Palliative care provision in PF is poor; there is limited use of advance care plans (ACP), high levels of end-stage healthcare utilisation, frequent hospital deaths and poor symptom control (6, 7). PF has a significant social impact and can disrupt relationships with close others/carers (8, 9).

There is consensus that earlier palliative care input is beneficial, with unpaid/family carer support and experienced healthcare professional involvement as important, but the model of care required is not clear. Healthcare professionals reportedly experience uncertainty about symptom management and the benefits of interventions(9).

Barriers to palliative care delivery have been identified as delays in palliative care referrals and end-of-life decisions, patients’ fear of talking about the future, prognostic uncertainty, confusion about the role of palliative care, discrepancies in understanding and care goals among patients, family, and medical staff, and inadequate communication about goals of care (10, 11). PF symptom burden is high despite available evidence-based interventions, guidelines, and information resources. The tailoring of existing evidence, knowledge and methods to produce the optimal model of palliative care for people living with pulmonary fibrosis in the UK has not been realised.

### Aim

This study aimed to establish patient views on how palliative care services and approaches can better meet the needs of people living with pulmonary fibrosis.

#### Research Question

1. What are palliative care and support needs in pulmonary fibrosis?
2. What are the barriers and facilitators to palliative care for people living with pulmonary fibrosis?
3. What existing supportive/palliative interventions, or components of these interventions, are acceptable to people living with pulmonary fibrosis?

## Methods

### Design

An exploratory qualitative approach was adopted using primarily focus group methodology to gain a range of views, stimulate discussion and prompt reflection. Focus groups encourage group interaction, generating discussion by allowing participants to raise issues of importance to them individually and explore them as a group, thus generating new ideas and questions, and highlighting any concerns and priorities (12). Semi-structured interviews were conducted with those who did not feel able to contribute in a group setting. Patients with PF were consulted on study design and informed the topic guide development. The Consolidated Criteria for Reporting Qualitative Research guidelines were used(13).

### Study Setting

The study was conducted between March–August 2024 within one National Health Service (NHS) Trust in England.

## Population

### Recruitment and Sampling

Participants were recruited via convenience sampling. Potential participants who met the inclusion and exclusion criteria were identified from clinic lists of patients by the clinical team (Table 1).

**Table 1:**
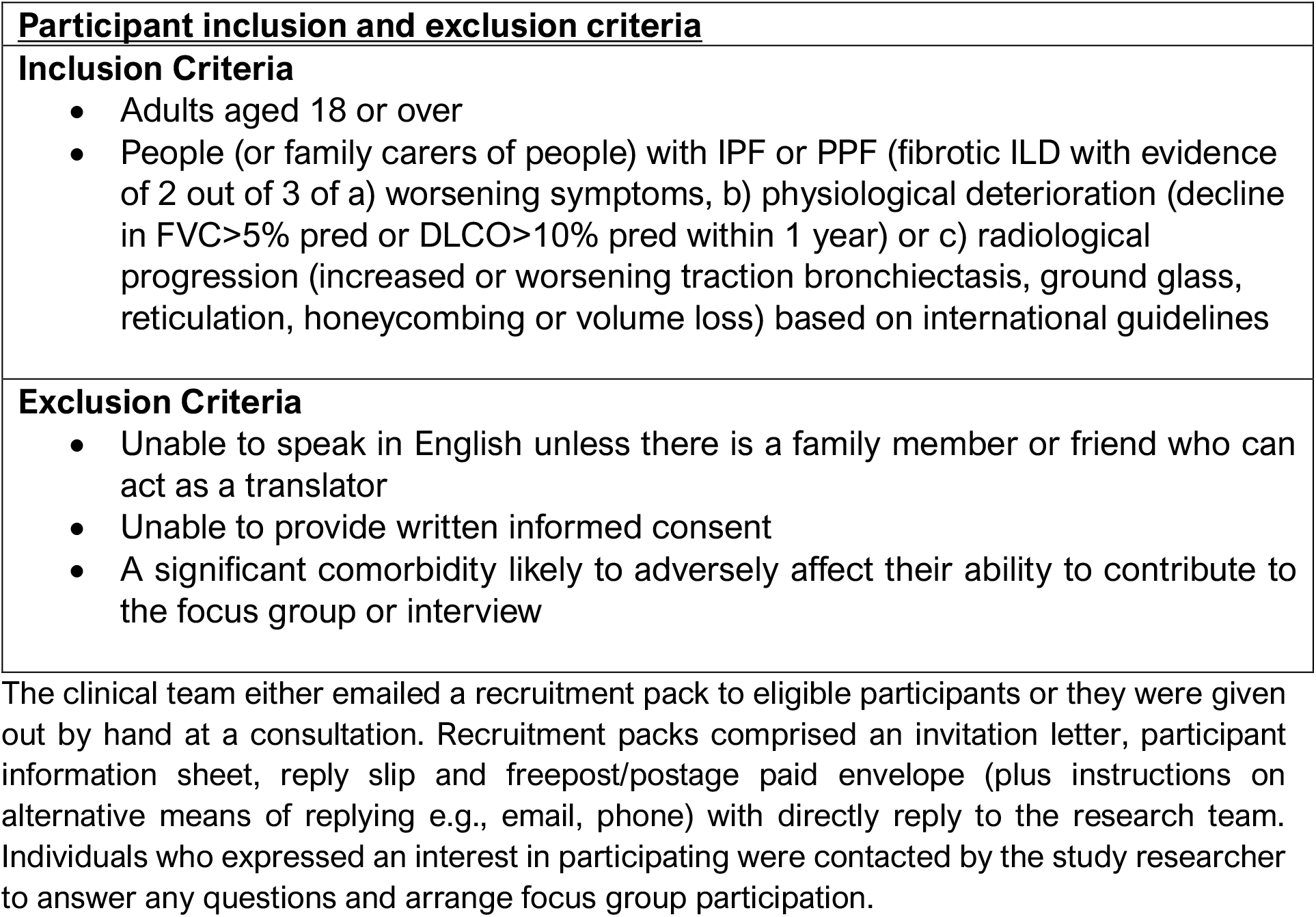
Inclusion and Exclusion Criteria.

| **Participant inclusion and exclusion criteria** |
| --- |
| **Inclusion Criteria**<br>• Adults aged 18 or over<br>• People (or family carers of people) with IPF or PPF (fibrotic ILD with evidence of 2 out of 3 of a) worsening symptoms, b) physiological deterioration (decline in FVC > 5% pred or DLCO > 10% pred within 1 year) or c) radiological progression (increased or worsening traction bronchiectasis, ground glass, reticulation, honeycombing or volume loss) based on international guidelines |
| **Exclusion Criteria**<br>• Unable to speak in English unless there is a family member or friend who can act as a translator<br>• Unable to provide written informed consent<br>• A significant comorbidity likely to adversely affect their ability to contribute to the focus group or interview |

The clinical team either emailed a recruitment pack to eligible participants or they were given out by hand at a consultation. Recruitment packs comprised an invitation letter, participant information sheet, reply slip and freepost/postage paid envelope (plus instructions on alternative means of replying e.g., email, phone) with directly reply to the research team. Individuals who expressed an interest in participating were contacted by the study researcher to answer any questions and arrange focus group participation.

### Data Collection and Analysis

Two focus groups and four interviews were conducted involving nine participants. One participant was accompanied by a family carer. One of the focus groups was held in person and the second was held online, in line with participant preferences. All interviews were undertaken on-line/via telephone. The focus groups lasted approximately ninety minutes and the interviews between 30-45 minutes. Focused groups and interviews were conducted by a senior research associate with several years’ experience in qualitative research and palliative care. (CG). The researcher was not known to the participants prior to the data collection Participants were made aware of the researcher’s background and independence from the clinical team.

Participants completed a demographic questionnaire and were then asked to discuss topic guided questions (table 2). Focus groups and interviews were audio recorded with permission, (except one interview where permission to record was refused, and notes were taken). At the end of the focus groups/interviews participants were given debrief sheets providing contact details for any required after support via a specialist nurse.

**Table 2:** Focus Group Schedule.

|  |
| --- |
| **(1) Understanding Need** |
| a) Can you briefly share with us how long you have been living with pulmonary fibrosis and what sorts of challenges it brings? |
| b) Are there any areas where you have needed more support (NB use diagram if needed as prompt) |
| c) What service, support or professional have you found most helpful for these needs? |
| d) What has not been less helpful, or could be improved upon? |
| **(2) Understanding of Palliative Care** |
| a) Have you heard of "palliative care"? Prompt: What does it mean to you? What do you think it means to other people? Does it mean anything else [aside from end of life, if mentioned]? |
| b) *Definitions: We mean palliative care to be support and treatment intended to improve quality the quality of life, such as helping people with their symptoms rather than treating the underlying disease itself. End of life care is often used to describe the last year of life* |
| c) What do you think the aim of palliative care might be in pulmonary fibrosis? (use prompt diagram 3 if needed) And what might its benefits? |
| d) What palliative care services or support are you aware of for people living with pulmonary fibrosis? [PROMPT – for patients and for families/carers] |
| e) Do you think that there are specific needs for people with pulmonary fibrosis which might be different from other conditions? |
| f) When do you think is the right time to start talking about how palliative care can help people living with pulmonary fibrosis? (see trajectory diagram) |
| g) Have you spoken about your wishes and preferences for care and treatment as you become more unwell? |
| h) Have you had all the information and preparation you would like about what the future might bring in this condition? – additional question if needed |
| i) Exit question: Is there anything else you would like to say about support you have received ? |

Audio recordings were fully transcribed, checked for accuracy and anonymised. Transcripts were read for familiarisation and conventional content analysis conducted(14). An initial framework was developed by CG from key issues arising from both the data and the topic guide. A subsequent coding framework was agreed by all co-authors to guide further analysis, organise the data, identify data saturation and develop the final themes.

## Results

### Participants Characteristics

Table 3 summarises the patient characteristics, relevant treatments, and self-reported participation in future care planning. The majority of participants were male. Demographic data provided was incomplete for two participants.

**Table 3:** Participant Characteristics.

| **Sample Characteristics** |  | **% (n)** |
| --- | --- | --- |
| **Gender** | Female | 11 (1) |
|  | Male | 89 (8) |
| **Age Range** | 50-59 | 11 (1) |
|  | 60-69 | 0 |
|  | 70- 79 | 67 (6) |
|  | 80 -89 | 22 (2) |
| **Occupation** | Engineer | 33 (3) |
|  | Teacher | 11 (1) |
|  | Sales | 11 (1) |
|  | Doctor | 11 (1) |
|  | Managing Director | 11 (1) |
|  | Self Employed | 11 (1) |
|  | Musician | 11 (1) |
| **Treatment and Support Received** | Antifibrotic Therapy | 44 (4) |
|  | Pulmonary Rehabilitation | 67 (6) |
|  | Peer Support Group | 44 (4) |
|  | Oxygen Therapy | 67 (6) |
|  | Morphine/Oramorph or Oxycodone | 0 |
|  | Breathing Exercises/Training | 4 (4) |
|  | Community Matron | 0 |
|  | Palliative Care | 22 (2) |
|  | Carers Assessment | 0 |
| **Participation in Future Care Planning** | ReSPECT form | 11 (1) |
|  | Advance Decision to Refuse Treatment | 22 (2) |
|  | Lasting Power of Attorney | 33 (3) |
|  | Living Will | 11 (1) |
|  | Advance Care Planning | 1 (1) |

### Findings

Four overarching themes were identified relating to palliative care approaches which could better meet the needs of this group: (1) Mechanisms for identifying and enacting patient palliative care needs; (2) Open channels of communication with professionals; (3) Access to palliative care across respiratory, community and specialist services and; (4) support for family carers.

#### 1. Mechanisms for identifying and enacting patient palliative care needs

Palliative needs within the group varied. Differences included information needs about future care planning, symptom management and whether support for day-to-day activities was wanted. Key to addressing these differences was the adoption of person-centred principles with discussions taking place when patients were ready and led by their priorities. This was exemplified by discussions about the right time to discuss their prognosis, or future care needs, which one participant noted should be *“down to each patient”* and *“when people need it”* (FG1-01).

The importance of being an active participant in discussions about their needs was also emphasised:

“*I think that for me personally I would like to be involved in anything that is going to be used for me, so medication or the different way of handling the condition I would definitely like to be informed and then I can make up my mind because it would be an informed decision rather than just guessing whether it should or shouldn’t be done*.*”* (FG2-04)

*“(The focus should be on) providing scope for the patient to have some input… rather than the professionals* (FG1-01)

Experiences of a person-centred approach also varied. Some reflected on occasions when healthcare professionals had involved them in shared decision making, whilst others were unsure who they should talk to about certain issues or how to contact appropriate services. It was also observed more generally that some patients can lack confidence to initiate, or contribute to, conversations about the care and support they need, and there was a need for services to proactively create opportunities to enable the patient to be *“an active participant in that process”* (FG1-C1).

*“Yes, I definitely think that because we’re all different …I’m pretty outgoing and I speak my mind where somebody might be really timid and they feel, ‘I can’t really ask that question. I need some help with the housework but I’m a bit embarrassed because maybe people will think I’m too lazy and I’m just being a hypochondriac and I just want to get help when I don’t really need it*.*’ So (something proactive) I think would be really helpful”* (FG2-04).

Patients suggested that checklists or information about the anticipated trajectory of pulmonary fibrosis could help them consider whether *“this didn’t happen, this has happened, this is worse, this isn’t”* (FG1-02), assist patients to *“put forward their own ideas*” (FG1-01) and give *“the person on the other side (healthcare professionals) somewhere to start from”* (P1-01).

The value of some kind of checklist that could help people identify support needed, or supportive in-put required in response to need, was also highlighted.

*“I think maybe if you had a list of the different services and what they pertained to and contact details, then it would be easier for either the carer or the patient to say, ‘Okay, well, I need more support with cleaning the house and that, so I need to contact so and so and they’ll be able to direct me to getting benefits for this or that,’ etc. because if you don’t know what you can get, how do you know what to ask for?”* (FG2-04).

#### 2. Open channels of communication with professionals

The participants described being actively involved in managing the impacts of their condition, such as infection and oxygen management. However, they also observed that this brought challenges with which they needed support, such as understanding the meaning behind changes in their symptoms and knowing who was the right person to contact for support.

To address this, readily available channels of communication with the professionals was seen as important; particularly the ability to initiate contact with professionals outside of scheduled consultations. Some already had access to email or a telephone number they could ring, when necessary, with one participant noting positively *“if I ever need anything, within the day they’ve (a healthcare professional) phoned me back”* (FG1-02).

Others identified that it would be useful to have “*a phone number that anyone could access”* (FG2-03) and “*someone on the end of the phone at the time when you think you really need them to discuss the problem you’ve got. Not to say ‘we’ll get in touch with you’*” (FG1-C1). Attention was also drawn to the importance such a service could play as their condition advanced: *“What other care is required down the track?, where more support is required?, how is that going? – it’s back to having somebody at the end of the phone”* (FG1-01).

The participants also felt that this was a service that could be provided by a range of professionals including respiratory specialists, primary care professionals or someone with a non-clinical role. One participant noted:

“*it may be useful to have a member of the admin staff in the respiratory (department) for example, that does know things, … if I do start to want to get a disabled pass to park somewhere can I talk to someone at the hospital that’s maybe done a little bit of research into it, that knows what I should be doing?…Maybe (someone who can provide) assistance with the day-to-day self-management of the disease from parking, to support, to finance, to whatever it is. That probably would be very useful”* (FG2-02).

Others suggested that it would also be useful to have a list of numbers from various services that they could contact as necessary.

Proactive contact from healthcare professionals either by phone or via a key contact in the community was also seen as potentially useful. One participant reflected that *“it would be nice to have somebody just contact maybe once in a while to see how I’m getting on”* (PI-02). Another felt that regular meetings with a community-based professional could be useful for considering holistic needs and discussing relevant responses.

#### 3) Access to palliative care across respiratory, community and specialist services

Participants described how, in the earlier stages of their condition, palliative care interventions took place primarily within respiratory care settings and via community groups such as peer support groups. Participants appreciated being able to discuss issues with known clinicians, and having a range of options for finding out those areas where they may need support and what might help (e.g. via other patients and newsletters).

Respiratory care was also seen as an appropriate starting point for conversations about disease progression and future planning. This was exemplified by two of the participants who were actively considering their future care needs. One had recently had a conversation with a respiratory specialist about prognosis and a second outlined his plans to use his next respiratory appointment to discuss “*What are the next steps?*… *What’s likely to happen?”* (PI-01).

There were, however, mixed views regarding expectations and preferences about who would be the main contact for palliative care as their condition advanced. Many hoped and anticipated that palliative care would continue to be delivered by the respiratory services, including home-based care. One participant commented that *“My wish would be that the (current respiratory team) follow me up to the end really”* (PI-01) whereas another felt it should be “*a bit like the respiratory support we get at the moment but extending that into whatever care is required down the track*” (FG-01). Others emphasised the potential role for community services such as primary care, community nurse and palliative care specialists. This, they felt, would enable easier access as their health declined. However, there was also some apprehension based on existing experiences relating to difficulties in accessing primary care and the extent to which community and primary care had the level of specialist knowledge required to offer the necessary support.

There were also contradictory views about the potential role of specialist palliative care. Many had little understanding of this service with one patient reflecting “*I really know next to nothing about palliative care to be honest. I suppose I’m just hoping it will be there”* (PI-01). Two participants, who had had some contact and knowledge with these services, expressed the view that they had been referred prematurely and preferred to be supported within respiratory services. However one participant, who had seen first-hand the role of specialist palliative care in supporting a family member, was clear about the need for a similar service for PF, arguing that *“the main thing…is a contact point that starts when you’re told you’re end of life”* suggesting more specifically this should involve “*a nurse who’s assigned to you… somebody who is end of life”* (PI-03). Drawing on the experiences of his family member he highlighted further how having a “*phone number of (someone in palliative care services) would be good to understand what’s happening and have an initial consultation and understand what is going to happen and what’s going to be offered”* (PI-03).

More generally, many participants observed that the services involved in supporting people with pulmonary fibrosis could be more streamlined and co-ordinated (e.g. provide faster communication of test results to patients and across settings). This was seen to become more pertinent as patients moved towards end of life.

*“Everything takes so long, you know, to get to the point where the next bit of treatment goes. And I haven’t worried too much until now, but when your life has a conclusion, you’d like the treatment to happen a little bit quicker* “(PI-03)

#### 4. Support for Family Carers

Family members and friends were variously described as taking on a carer role through monitoring symptoms, identifying the need for additional help, support with practical tasks and providing emotional support.

*“My wife does tell me to breath*…*‘You’ve got to stop, and you’ve got to sit down’”*

(FG1-01)

There were mixed views from patients as to how far these carers were seen as in need of support. Most felt carers would need more support as the illness progressed. Discussing his partner, one participant noted “*We need to have an understanding of what the next few steps are. So, yeah. I think she needs some … she needs to be there, being given advice as well, I think”* [PI-01]. The need for family members to be supported at the end-of-life was noted so that they would know what to expect and who to contact. However, one patient also expressed reservations about his partner knowing more about his condition than he was willing to share and was therefore unsure about carers being involved or having their own sources of support.

## Discussion

### Main Findings of the Study

This study aimed to establish the palliative care needs of people with PF, and to understand the palliative care approach that might best meets these. Whilst there was divergence of views, a preference emerged for (1) open, accessible and responsive services, that (2) included mechanisms for both identifying and enacting the palliative needs, (3) across the whole respiratory pathway of both patients, and (4) their carers. These finding are consistent with, and add to, previous studies in this area.

A recent scoping review of the unmet needs of people living with IPF identified a need for better understanding of disease trajectory to aid in ACP(15). Supporting patients’ information needs has been identified as being important in addressing the low levels of engagement with ACP which is often found across many chronic respiratory diseases(16). Active participation with ACP is likely to require a proactive and open approach by respiratory clinicians, as people with pulmonary fibrosis have indicated a strong preference for conversations about end of life care earlier in their disease process, whilst still well enough to participate fully(17, 18). Identification of trigger points and appropriate training in ACP may support clinicians to meet this important need in pulmonary fibrosis(16, 19).

Patients were clear on their need for person-centred approaches to their care, and the identification and addressing of their support needs. This proactive approach could be achieved via the use of tools such as checklists, questionnaires, or information about the anticipated trajectory of pulmonary fibrosis. The Needs Assessment Tool-progressive disease: interstitial lung disease is a tool to help health professionals assess the needs of patient with ILD that is recommended by NICE(20).

Another intervention that could enable this is the Support Needs Approach for Patients (SNAP: https://thesnap.org.uk/)(21) which provides patients with a booklet containing an evidence-based set of questions to consider in order to help them identify their support needs that then informs a needs-led conversation with the health care professional. This intervention model maps well to patients’ desire for some kind of checklist that could help people identify support needed and supportive in-put required in response to need.

Our second finding, of the importance of open channels of communication for people living with PF, mirrors the patient preferences reported in a systematic review of participants with a range of palliative care conditions(22). In this study, effective palliative care was characterised by a triad of responsive healthcare professionals, responsive care environments and responsive palliative care organisation(22). In the ADAPT randomised controlled trial, which included patients with pulmonary fibrosis, phone called based social and psychological support improved quality of life over a six-month period(23). Unsurprisingly, given our findings, patients tend to be positive about their experiences of dedicated palliative care out of hours advice lines, although their effectiveness for the pulmonary fibrosis population is not known (24).

Our third finding was the importance of palliative care needs being met across palliative, respiratory and primary care services. NICE guidelines recommends that an ILD specialist nurse should be available “at all stages of the pathway” to provide information and support to people with IPF, their families and their carers (NICE CG163, 2013, updated 2017)(25). Many participants in this study saw respiratory care as an appropriate place to start discussions surrounding palliative care, however there were concerns about who would continue to be their main contact for palliative care as their disease progressed. Whilst relational factors are critical to meeting care needs, there is an observed tension between who might be best able to provide continuity, versus the level of specialist knowledge and skills needed to optimally manage the condition as it progresses(26, 27). Interestingly, the majority of ILD specialist nurses feel that they should be responsible for providing palliative care for their patients throughout the whole time-course of their disease(28).

Finally, support for unpaid/family carers was identified as an important component of palliative care, and care from respiratory physicians and specialist nurses, although there were mixed views about how best to meet those needs. Whilst the need for general support for carers by palliative care is identified in NICE guidance (CG163, 2013, updated 2017)(25), the potential for varying or conflicting information needs, need for carer training and/or specific practical considerations are not addressed in this guidance. In this respect, a questionnaire study has recently reported that patients with pulmonary fibrosis have different information needs from their family carers, with patients having more needs related to ACP whereas cares wanted to know more about care provision at home(29). In previous pulmonary fibrosis studies (not specific to palliative care), carers have felt their role in clinical consultations to be ambiguous or passive, which adds to the challenges of accessing or processing information(30). Whilst we did not speak with carers directly in this study (other than one carer who accompanied a patient to a focus group), we would suggest further research is undertaken to understand the extent to which carers are willing and supported to feel actively involved in care decisions.

### What this study adds

Contributing to the current evidence base, people living with pulmonary fibrosis have identified service specific preferences around openness and responsiveness, and expressed a desire for a person-centred system that allows them to take a proactive role in their own care. Less consensus was reached on which healthcare professionals or service should be responsible for palliative care. It was identified that particular challenges are likely to arise when information needs in a patient-carer dyad are divergent, and for this reason an individualised, relational approach to palliative care is particularly important with complimentary support for unpaid carers

### Implications for Practice

Whilst some participants could see the value of palliative care services, participants were largely ambivalent as to *who* should provide such care, as long as their self-identified needs were met. Many participants recognised that care needs that might be considered palliative care already been addressed within their existing respiratory team infra-structure. Consideration must be given to whether specialist palliative care involvement or “ownership” of pulmonary fibrosis is an appropriate model of care in this population, and whether more work could be done to “in-reach” into ILD services to enhance the level of palliative care provided.

### Implications for Research

There is a lack of clinical trials into the efficacy, acceptability and cost effectiveness of interventions that aim to meet the palliative care needs of people living with pulmonary fibrosis. Given the importance placed on this approach by our participants, we would suggest that the utility of palliative care interventions in PF are explored further in UK based clinical trials.

### Limitations

This study was based within a single ILD service, with the possibility that findings were specific to this setting and not generalisable in a wider sense. Participants were largely male, and white British, and service recommendations would ideally be informed by a wider range of genders and ethnic diversity. Whilst participants living with pulmonary fibrosis identified carer support as important, the carers voice was not represented fully in this study and requires further exploration.

## Conclusions

More work is needed to understand the optimal approach to palliative care for people with pulmonary fibrosis. Future studies should aim to understand the impact of remote palliative care interventions that promote self-, as these have been identified as acceptable and potentially beneficial by our participants.

## Data Availability

All data produced in the present study are available upon reasonable request to the authors

## Acknowledgements

CB, MF and CG are funded by the National Institute for Health and Care Research (NIHR) Applied Research Collaboration East of England (NIHR ARC EoE) at Cambridgeshire and Peterborough NHS Foundation Trust. The views expressed are those of the author[s] and not necessarily those of the NIHR or the Department of Health and Social Care.

The study team wish to thank all the participants of the study, and the patients who helped inform the study design and topic guide.

## Ethics Declaration

Ethical approval was obtained from Wales Research Ethics Committee 6, reference 23/WA/0303, 16^th^ November 2023. IRAS project ID: 330149. Written informed consent was obtained from all participants.

## Contributions

AW and CB conceptualised the study. AW, CB and MF designed the study and wrote the protocol. CG undertook the interviews and qualitative analysis. CB and AW wrote the manuscript with contributions from MF, CG and IK. All authors critically analysed and approved the final version of the manuscript.

